# The role of the *ADRB2* Thr164Ile variant in lung function determination, plasma proteome variability and other phenotypes in UK Biobank

**DOI:** 10.1101/2024.10.14.24315217

**Authors:** Katherine A Fawcett, Robert J Hall, Richard Packer, Kayesha Coley, Nick Shrine, Louise V Wain, Martin D Tobin, Ian P Hall

## Abstract

**Introduction:** The effect of coding polymorphisms of the beta-2 adrenergic receptor gene (*ADRB2*) on functional properties of the receptor is well-established. We recently reported a genome-wide significant association between Thr164Ile and lung function, but the contribution of this variant to other traits remains unclear.

**Methods:** To identify pleiotropic effects of *ADRB2* Thr164Ile and other coding variants, we performed respiratory-focused and phenome-wide association studies in UK Biobank. In addition, we used available Olink proteomic data to characterise enriched pathways and upstream regulators of proteins associated with *ADRB2* polymorphisms.

**Results:** The minor allele of Thr164Ile was associated with reduced lung function, but not COPD or asthma defined using self-report and diagnostic codes in healthcare records. It was also associated with non-respiratory traits including increased eosinophil counts and blood lipid measurements, including increased cholesterol, reduced triglycerides and reduced apolipoprotein A. Proteins associated with Thr164Ile (P-value≤0.01) were enriched for various pathways, with the eosinophil-raising allele associated with reduced neutrophil degranulation, immunoregulatory interactions between a Lymphoid and a non-Lymphoid cells, TNF binding and DAP12 interactions, as well as activation of lipid metabolism pathways, including FXR/RXR activation and LXR/RXR activation. A gene-based analysis of rare, non-synonymous *ADRB2* variants, identified a novel association with non-rheumatic pulmonary valve disorders, but no association with lung function.

**Discussion:** In conclusion, the lung function-lowering allele of Thr164Ile is associated with traits and proteins indicative of a role in immune and lipid metabolism pathways, but not COPD or asthma. In contrast, *ADRB2* rare coding variants are not associated with lung function.

## Introduction

Three coding region polymorphisms in the *ADRB2* gene are known to alter the behaviour of the beta 2 adrenoceptor (ADRB2) in recombinant cell lines [1, 2] and in ex vivo human airway myofibroblasts in primary cell culture [1-3]. In brief, the Thr164Ile variant (rs1800888) alters signalling after receptor stimulation with a range of beta 2 adrenoceptor agonists, with reduced signalling for catechol ligands with the Ile164 form of the receptor [4]. The Arg16Gly (rs1042713) and Gln27Glu (rs1042714) variants alter receptor downregulation profiles following stimulation, with greater downregulation being observed with Gly16 and reduced downregulation with Glu27 [1-3].

There have been a large number of clinical studies which have explored the relevance of these polymorphisms to a range of respiratory and non-respiratory conditions, including asthma [5-7] and chronic obstructive pulmonary disease (COPD) [8-13]. Initial small studies in selected populations suggested that these polymorphisms are associated with asthma per se. However, the largest study to formally assess this to date, which used the UK 1958 birth cohort, showed no significant genotype-dependent effects on asthma risk [5]. Subsequently, there have been two systematic reviews addressing the contribution of these variants to asthma risk, which have come to differing conclusions [7, 14]. For COPD, a systematic review and meta-analysis of 16 studies found no association between the Thr164Ile, Arg16Gly and Gln27Glu variants and COPD but was underpowered to investigate the Thr164Ile variant due to its lower minor allele frequency (MAF) [10]. A Danish population-based study also tested all three variants and identified associations between Thr164Ile and lung function and COPD [12].

We recently described an independent, genome-wide association between the Thr164Ile variant and lung function in a multi-ancestry study including 588,452 individuals [15]. In order to gain a fuller understanding of the potential basis for this association, we used UK Biobank to explore association of the Thr164Ile variant with a broad range of respiratory and non-respiratory traits. In addition, by using the exome sequencing data from the same population, we explored the potential contribution of rare coding variants to these traits across multiple genetic ancestry groups. Finally, we explored associations between the Thr164Ile variant and the plasma proteome in the subset of UK Biobank with Olink data and describe here the pathways underlying the associated proteins.

## Methods

### Study population

The UK Biobank comprises half a million volunteer participants from the UK, aged 40-69 at recruitment. Baseline measurements, including spirometry-based lung function and anthropometric measurements, were taken at recruitment and at follow up periods. Lifestyle and medical history information was self-reported through questionnaires and in a nurse’s interview. Secondary care records (hospital episode statistics) are also available, and 45% of participants have linked primary health care records, including prescription records. Blood samples were taken at baseline for genome-wide genotyping and were later used for exome and whole-genome sequencing. Protein measurements for 2,923 proteins using the Olink platform were also recently released for 54,219 participants.

Genotyping was carried out using the Affymetrix Axiom® UK BiLEVE array or the Affymetrix Axiom® UK Biobank array. This data was used as a basis for assigning individuals to ancestry groups using ADMIXTURE v1.3.0 (see [15] for details) and for excluding related individuals. The Thr164Ile, Arg16Gly and Gln27Glu polymorphisms were directly genotyped in UK Biobank. Genotypes for rare variants in *ADRB2* were extracted from exome sequencing data. Exome capture was performed using the IDT xGen Exome Research Panel v1.0 including supplemental probes, and paired-end sequenced using the Illumina NovaSeq 6000 platform (see [16] for additional details). Variants were called using DeepVariant 0.0.10 and aggregated into gVCFs with GLnexus 1.2.6 using the default joint-genotyping parameters for DeepVariant.

### Phenome-wide association studies

We used DeepPheWAS [17] to test association between *ADRB2* coding variants and up to 1,908 clinically-relevant traits in up to 419,936 UK Biobank participants. Briefly, this software constructs phenotype tables comprising inverse normal transformed continuous traits measured at UK Biobank assessment centres or from primary care records, as well as binary traits based on self-report and/or diagnostic codes within linked healthcare records. Binary traits with fewer than 50 cases and quantitative traits measured in fewer than 100 individuals were excluded. Association testing was then performed using either linear regression (for quantitative traits) or logistic regression (for binary traits), adjusting for age, sex, genotyping array and the first ten ancestry-based principal components (PCs). The false discovery rate was set to 1%.

In order to test for association with rare *ADRB2* variants, gene-based tests burden tests were run using the exome sequencing data. Regenie v3.3 was used to create masks of predicted loss of function and missense variants within five different frequency categories: singleton variants, variants with minor allele frequency (MAF)<0.00001, MAF<0.0001, MAF<0.001, and MAF<0.01. Individuals were coded as 0 if they had no rare alleles across the *ADRB2* gene, 1 if they were heterozygous for at least one rare allele, and 2 if they were homozygous for at least one rare allele. For both single-variant and gene-based analyses, summary statistics were output to tables and results visualised in graphs using DeepPheWAS.

### Respiratory-focused phenome-wide association studies

We conducted a look-up of the Thr164Ile variant in 26 genome-wide association studies (GWAS) across ten clinical respiratory traits: asthma, bronchiectasis, bronchopneumonia, chronic bronchitis, chronic obstructive pulmonary disease (COPD), chronic sputum production, emphysema, idiopathic pulmonary fibrosis (IPF), respiratory infections, and interstitial lung abnormalities (ILA). Some of the GWAS summary statistics are publicly available (on GWAS Catalog/online storage) and some are available upon request (Supplementary Table 1).

### Smoking subgroup analyses

We tested associations with COPD and asthma in smoking subgroups using logistic regression models in never-smokers and ever-smokers (based on UK Biobank field 20160). For asthma, we included age, sex, genotyping array and ancestry-based PCs in the model and, for COPD, we also included standing height as a covariate.

### Multi-ancestry analyses

Given the lower frequency of the Thr164Ile variant in non-European ancestry clusters (MAF = 0.002, 0.000, and 0.005 in the African, East Asian and South Asian clusters respectively), we were underpowered to detect its effects on lung function in these groups (power = 10%, 0%, and 18% respectively). However, we extended our gene-based phenome-wide association study to investigate the effect of this variant along with other rare variants in individuals of African, East Asian and South Asian ancestry. Quanto v1.2.4 was used for power calculations.

### Protein analysis

Protein levels were measured from blood using the Olink platform in 54,219 UK Biobank participants. The latest release of this data includes 2,923 unique protein assays, not including ADRB2. We tested for association between Thr164Ile and untransformed log2 fold protein levels, assuming an additive genetic model and including covariates: olink batch, age, sex, genotyping array and the first ten genotyping-based PCs. All proteins showing association (P<0.01) were used as input to Qiagen Ingenuity Pathway Analysis (IPA), along with their p-value and beta value, to identify enriched pathways and potential upstream regulators. Colocalization was carried out using coloc.susie software [18] to ascertain whether both the Thr164Ile lung function signal and the associated protein signals were driven by the same variant, with regional summary statistics from lung function and protein analyses and a linkage disequilibrium reference panel based on 10,000 randomly selected European individuals from UK Biobank. All variants within a 2Mb region centred on the Thr164Ile variant and with a MAF > 0.001 were included in this analysis. We extended these analyses to Arg16Gly and Gln27Glu to ascertain overlap between proteins and pathways impacted by *ADRB2* coding polymorphism.

## Results

### Phenome-wide association studies

Given the genome-wide significant association we previously described between the *ADRB2* Thr164Ile variant [15] and lung function, we performed a PheWAS in UK Biobank in order to understand its clinical relevance across a broad range of respiratory and non-respiratory traits. As previously shown, the minor allele was associated with reduced lung function, including forced expiratory volume in 1 second (FEV_1_)/forced vital capacity (FVC) (p-value = 3.06×10^−19^) and peak expiratory flow (PEF) (p-value = 9.95×10^−10^). It was also associated with increased eosinophils, a variety of blood lipid measurements including increased cholesterol, reduced triglycerides and reduced apolipoprotein A, as well as reduced creatinine and hand grip strength. The only binary outcomes associated with Thr164Ile were spirometrically-defined COPD (GOLD1+) and hyperplasia of the prostate (Figure 1, full results in Supplementary Table 2). There was no significant association with COPD defined using self-report and diagnostic codes in electronic healthcare records (EHRs) in UK biobank (OR=1.01, CI 0.93-1.09, p-value = 0.873) or in a look-up of the Thr164Ile in 26 GWAS of respiratory diseases (Supplementary Figure 1 and Supplementary Table 3).

**Figure 1.**
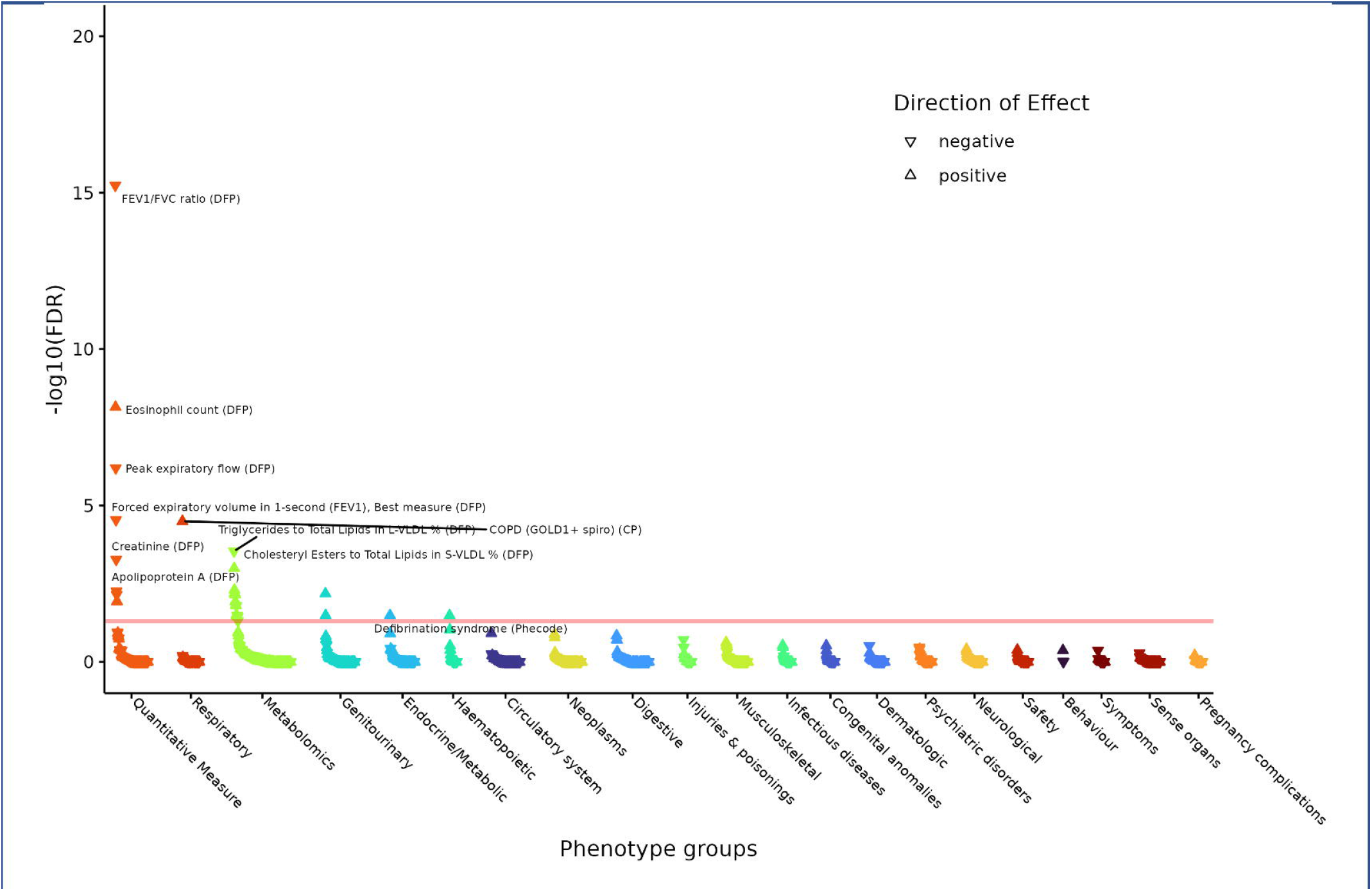
Phenome-wide association of UK Biobank traits with the Thr164Ile variant. The directions of effect are aligned with the minor T allele.

Due to previous reports that the effects of *ADRB2* coding variants can be modified by smoking status [13, 19-21], we also tested association of Thr164Ile with self-reported and EHR-coded COPD and asthma in UK Biobank never- and ever-smokers separately. No associations were found within these subgroups (Supplementary Table 4) and there was no evidence for an interactive effect between Thr164Ile and smoking on either COPD (p-value = 0.958) or asthma (p-value = 0.633).

Additionally, some prior studies have identified associations between *ADRB2* coding SNPs and COPD and asthma under different genetic models [7, 13]. We therefore tested Thr164Ile under a dominant and recessive model in UK Biobank but found no associations with either COPD or asthma (Supplementary Table 5), although given the low frequency of the Thr164Ile variant these studies are likely to be underpowered to detect a small effect.

### Proteins and pathways

For mechanistic insight into the effects of Thr164Ile on lung function, we tested its association with 2,923 proteins measured in UK Biobank blood samples (N = 54,219) using the Olink platform. Only three associations were discovered at a P value threshold corrected for the effective number of independent tests (as calculated by MeffLi method), which was with MEGF10 (Multiple Epidermal Growth Factor-Like Domains Protein 10) (p-value = 7.84×10^−7^), TNF superfamily member 13b (TNFSF13B) (p-value = 3.05×10^−5^), and T cell immunoglobulin and mucin domain containing 4 (TIMD4) (p-value=3.27×10^−5^).

At a nominal significance threshold (p-value<0.01), the Thr164Ile variant was associated with 61 proteins (Supplementary Table 6). These proteins were analysed in IPA to identify enriched canonical pathways and potential upstream regulators (Figure 2, Supplementary Tables 7 and 8). Thirty-seven pathways were enriched amongst proteins associated with the Thr164Ile minor allele (Supplementary Table 7). These included inflammatory response pathways such as neutrophil degranulation, immunoregulatory interactions between a lymphoid and a non-lymphoid cell, and TNFs binding to their receptors (predicted to be inhibited as a result of Thr164Ile-associated protein changes). There was also enrichment for some metabolic pathways, including FXR/RXR activation, LXR/RXR activation, hepatic cholestasis (inhibited), which are all involved in lipid metabolism. Finally, pathways associated with wound healing were also enriched, including hematoma resolution (activated). Predicted upstream regulators (Supplementary Table 8) were dominated by those involved in immune response including TNF, IL-2 and IL-1β, which were predicted to be inhibited upstream regulators, while hepatitis A virus cellular receptor 1 (HAVCR1) was predicted to be activated.

**Figure 2.**
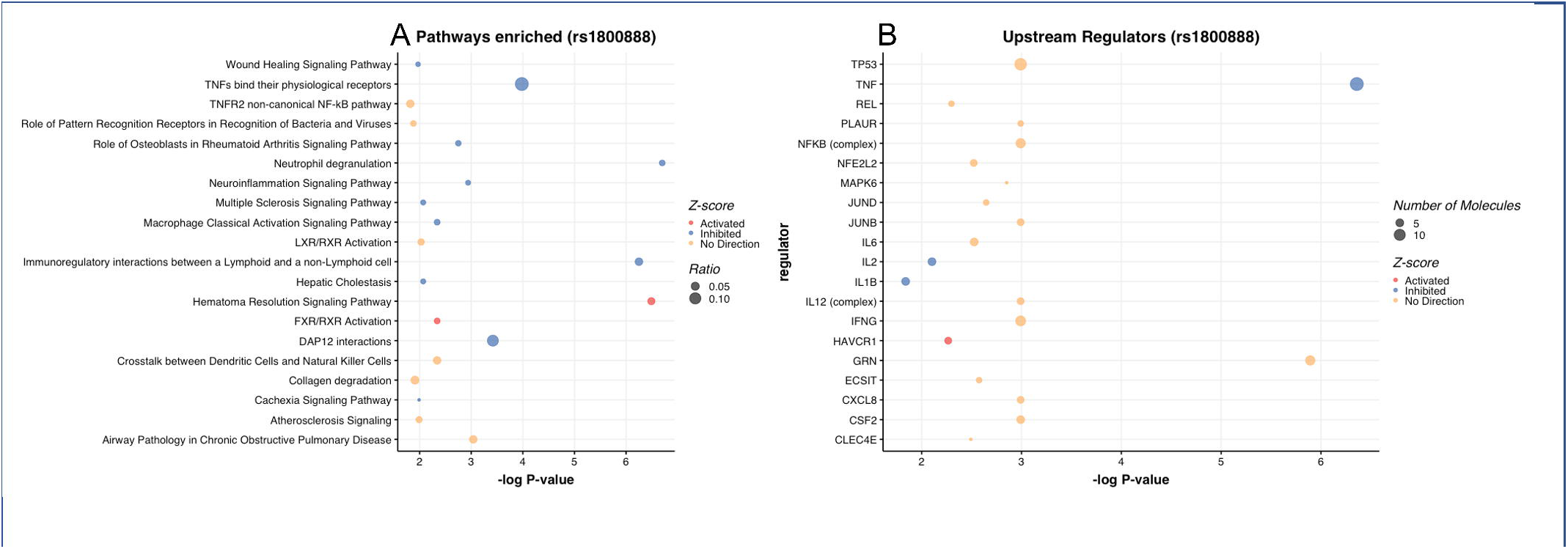
Bubble plots of enriched pathways and upstream regulators amongst proteins associated with the Thr164Ile variant, generated using Ingenuity Pathway Analysis. The colour of the circles indicates whether or not the pathway is predicted to be activated or inhibited (or whether no direction can be determined), and the size of the circles indicates the percentages of proteins in the full pathway that are amongst the Thr164Ile-associated proteins.

### Rare coding variants in ADRB2

Exome sequencing across the full UK Biobank cohort detected 264 rare (MAF<0.01) non-synonymous changes in *ADRB2* (Supplementary Table 9). To assess whether the effects of Thr164Ile on lung function might be mediated by rare variants or whether rare variants had independent effects on respiratory or non-respiratory traits in UK Biobank, we performed another PheWAS in individuals of European genetic ancestry. Given lack of statistical power to detect effects of single, rare variants, we collapsed these variants together using different maximum MAF cut-offs. Only one of these models (variants with MAF<0.0001) showed any associations at an FDR < 0.01: namely with nonrheumatic pulmonary valve disorders (OR = 11.497, CI 4.247-31.121, p-value = 1.53×10^−6^; Figure 3 and Supplementary Table 10).

**Figure 3.**
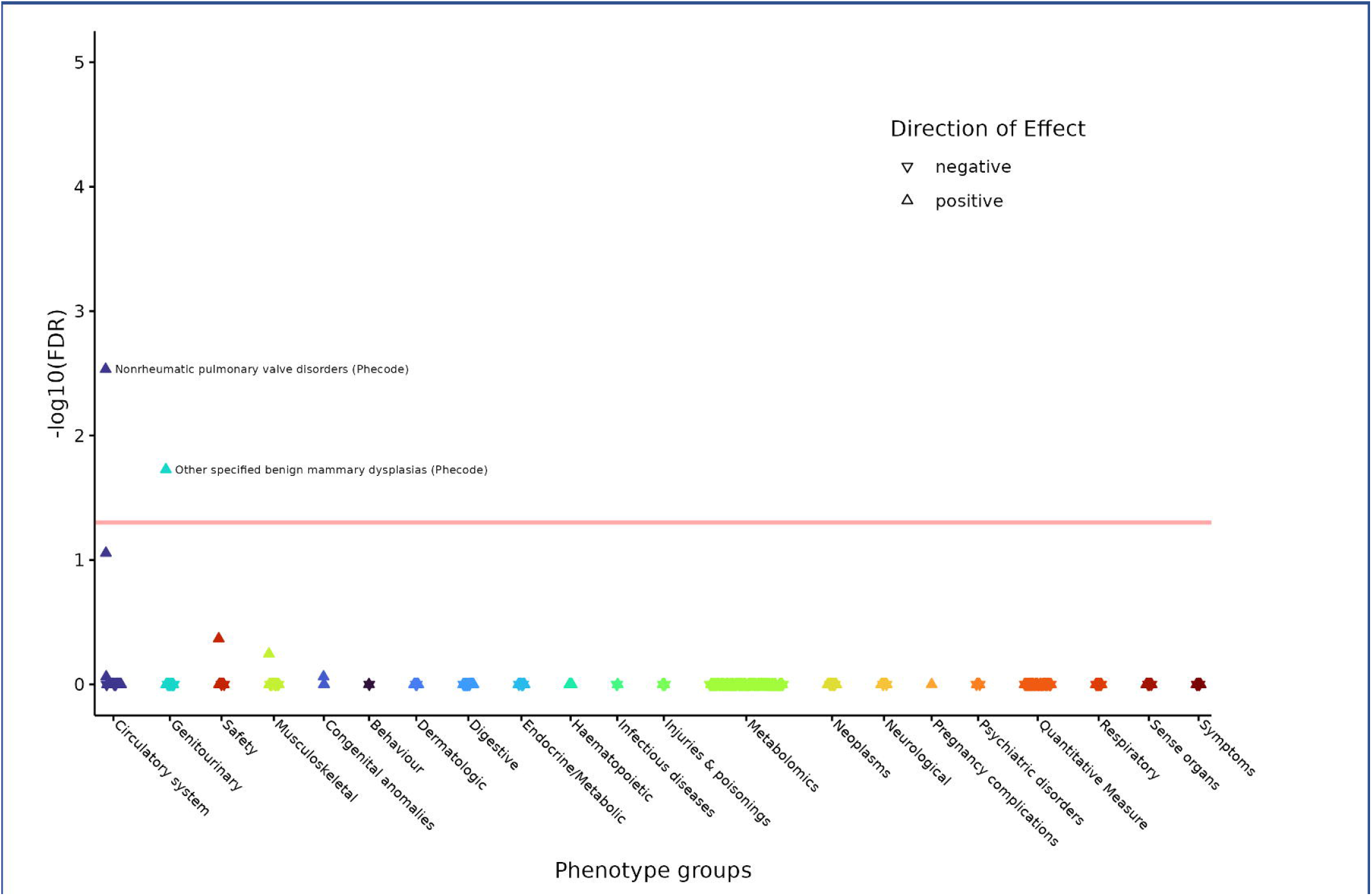
Phenome-wide association of UK Biobank traits with a gene-based model of rare (MAF<0.0001), non-synonymous *ADRB2* variants.

In UK Biobank participants within African, East Asian and South Asian genotyping-based clusters, there were no associations in *ADRB2* gene-based tests, even at a lenient FDR threshold of 5% (Supplementary Tables 11-13).

### Arg16Gly and Gln27Glu variants

The two other coding polymorphisms in *ADRB2* (Arg16Gly and Gln27Glu) have minor allele frequencies of 0.42 and 0.32 in UK Biobank respectively. They are in linkage disequilibrium (r^2^ = 0.43); the minor A allele of Arg16Gly tending to be inherited with the major C allele of Gln27Glu. In previous studies, they have shown inconsistent associations with a variety of respiratory conditions and associated traits. We therefore performed PheWAS of these SNPs and showed association with increased eosinophil count and decreased neutrophil count in the blood (Supplementary Figures 2A and B, full results in Supplementary Tables 14 and 15). They showed no association with respiratory conditions (Supplementary Table 3), including COPD or asthma in UK Biobank ever- and never-smokers and under different genetic models (Supplementary Tables 4 and 5 respectively).

In the UK Biobank Olink data, the Arg16Gly and Gln27Glu variants were associated with 11 and 22 proteins respectively, at an adjusted P value threshold (or −log10P = 4.47, highlighted red in Supplementary Tables 16 and 17) and with 76 and 123 proteins at a nominal P value threshold (P<0.01) respectively. We used IPA to identify enriched canonical pathways and potential upstream regulators (Supplementary Tables 18-21, Supplementary Figure 3). The results shown are aligned to the allele associated with higher eosinophil and lower neutrophil numbers. In brief, Arg16Gly-associated proteins were enriched in 17 pathways (Benjamini–Hochberg corrected p-value<0.05) including neutrophil degranulation, wound healing, extracellular matrix degradation, osteoblast function in rheumatoid arthritis (all predicted to be inhibited), and matrix metalloproteinase (MMP) activation and osteoarthritis (no directionality predicted). Gln27Glu-associated proteins were enriched in 51 pathways, including 12 that overlap with the Arg16Gly-enriched pathways as well as inflammatory pathways such as IL-17 signalling, Th1 and Th2 pathways, pathogen induced cytokine storm pathway (all predicted to be inhibited), and airway inflammation in asthma (no specific directionality predicted). We compared these results to the pathways enriched amongst Thr164Ile-associated proteins and found five overlapping pathways between all three coding variants (Supplementary Figure 4A): neutrophil degranulation (inhibited), wound healing signalling pathway (inhibited), activation of matrix metalloproteinases (no direction), airway pathology in chronic obstructive pulmonary disease and role of osteoblasts in rheumatoid arthritis signalling pathway (inhibited). Analyses of upstream regulators in general showed no clear directional effects although for Arg16Gly Tumour necrosis factor (TNF) and interleukin-17A (IL-17A) were both predicted to be inhibited upstream regulators, whilst for Gln27Glu predicted inhibited upstream regulators included TNF, IL-18 and c-Jun NH2-terminal kinase (JNK). Overlap of upstream regulators between all three variants is shown in Supplementary Figure 4B.

## Discussion

Here, in comprehensive PheWAS analyses, we confirm that the Thr164Ile *ADRB2* variant is associated at a genome-wide significant level with lung function in UK Biobank, and in addition show the minor allele is associated with a range of non-respiratory phenotypes including eosinophil counts, blood lipid measurements as well as reduced creatinine and hand grip strength. Rare variant analyses, using a collapsing gene-based approach, identified an association with nonrheumatic pulmonary valve disorders. However, neither rare *ADRB2* variants nor the common *ADRB2* codon 16 and 27 polymorphisms showed association with lung function, COPD, asthma or any other respiratory diseases.

Previous smaller association studies exploring *ADRB2* coding region polymorphisms and asthma risk have yielded inconsistent results, exemplified by two systematic reviews [7, 14]; one concluding that Arg16Gly and Gln27Glu polymorphisms are associated with asthma risk, while the other found no evidence for association. Our study provides a definitive answer to this issue. The association of Thr164Ile with reduced lung function in UK Biobank is consistent with the results of a smaller Danish population-based study [12], which found association between Thr164Ile and reduced lung function and increased risk of COPD.

ADRB2 signals predominantly through elevation of intracellular cyclic AMP and protein kinase A activation. ADRB2 stimulation is known to cause many downstream effects including altered gene expression in a wide range of cell types driven through cyclic AMP response element binding protein (CREB) family members binding to CRE regulatory sites (reviewed in [22]). Given the PheWAS associations we observed between the Thr164Ile variant and lung function, eosinophil count and lipid measurements, we explored potential underlying mechanisms using the Olink plasma proteome data in a subset of UK Biobank. Amongst 61 proteins nominally associated with Thr164Ile, there were a number of enriched pathways and upstream regulators. Interestingly, the eosinophil-increasing allele was associated with protein changes indicative of inhibition of immune pathways including neutrophil degranulation, immunoregulatory interactions between a Lymphoid and a non-Lymphoid cells, TNF binding and DAP12 interactions amongst others. Consistent with its association with blood lipid measurements, it was also associated with pathways relevant to lipid metabolism including FXR/RXR activation, LXR/RXR activation and Hepatic Cholestasis. Interestingly, both exogenous and endogenous lipids are known to play a role in immune regulation, including immune cell activation, differentiation and expansion, suggesting possible crosstalk between these pathways and a role in immune-related traits and diseases [23].

Probably on account of its low frequency, only three proteins were associated with Thr164Ile at a P value threshold adjusted for multiple testing (MEGF10, TNFSF13B and TIMD4). Although speculative, the association of the lung function-lowering allele with lower MEGF10 levels suggests that this protein could have a role in maintaining airway calibre. MEGF10 is a member of the multiple epidermal growth factor-like domains protein family, and plays a role in cell adhesion, motility and proliferation. It is also an essential factor in the regulation of myogenesis where it is believed to control the balance between skeletal muscle satellite cell proliferation and differentiation through regulation of the notch signaling pathway. Mutations in *MEGF10* are associated with myopathies, and so it is possible that lower MEGF10 alters airway wall morphology through effects on airway myofibroblasts leading to the observed FEV_1_/FVC signal in our study [24]. In a look up of in house spatial transcriptomic data using fixed human lung tissue, *MEGF10* transcripts were detected in airway smooth muscle *in situ* (data not shown). Interestingly, myopathy caused by *MEGF10* mutation in a range of non-human models can be reversed by selective serotonin reuptake inhibitors (SSRIs) [25], so if reduction in MEGF10 function is causally related to lung function there is a possible route to drug repurposing.

An alternative explanation is that this pQTL association could have arisen due to an indirect effect of Thr164Ile on regulation of MEGF10. For example, because Thr164Ile is also associated with eosinophil count, proteins expressed in eosinophils could underlie the observed association. However, according to the Human Protein Atlas, MEGF10 is not detected in immune blood cells. A search of the STRING, IntAct, BioGRID and the Molecular INTeraction databases revealed no known interactions between ADRB2 and MEGF10.

The other two proteins associated with Thr164Ile, TNFS13B and TIMD4, have known roles in immune regulation. TNFSF13B is a cytokine with a role in proliferation, differentiation and activation of B cells and polymorphisms in this gene have been associated with autoimmune conditions [26]. TIMD4 is a phosphatidylserine receptor with roles in clearance of apoptotic cells and T cell regulation, including promotion of type 2 inflammation [27].

One interesting observation from our results is the dissocation of the genetic association signals for eosinophila and asthma risk at the *ADRB2* locus. Previous studies have generally shown substantial overlap between the genetic associations for these two phenotypes [28], and there is considerable evidence that eosinophilia is associated with poorly controlled asthma and treatment response in COPD [29, 30]. The dissociation we observed between eosinophil and asthma phenotypes implies that not all genetic factors involved in control of eosinophil levels lead inevitably to asthma.

This study had several limitations. Firstly, we only investigated nonsynonymous coding region variants, rather than synonymous variants, or intronic or upstream/downstream variants that might have roles in regulation of *ADRB2* splicing or expression. Secondly, our analysis of the Thr164Ile variant under a recessive genetic model will have been underpowered due to there being only 123 individuals carrying two copies of the minor allele in UK Biobank. Thirdly, most of our analyses were limited to the largest ancestry group in UK Biobank, namely white British individuals. A multi-ancestry analysis of *ADRB2* variants in additional, diverse cohorts could test generalisability of these findings to other world populations.

Taken together these results show that it is the Thr164Ile variant of *ADRB2* drives the lung function genetic association signal at this locus potentially through alteration of immune- and lipid-metabolism pathways.

## Data Availability

All summary statistics produced in the present work are contained in the manuscript. Researchers can apply for access to individual-level data from UK Biobank.

## Ethics statement

UK Biobank participants provided informed written consent and gained ethical approval from the North West Multi-center Research Ethics Committee, the National Information Governance Board for Health and Social Care in England and Wales, and the Community Health Index Advisory Group in Scotland (http://www.ukbiobank.ac.uk/ethics/). These specific analyses have been approved as part of UK Biobank projects 648, 56607, and 43027.

## Funding and acknowledgements

This research has been conducted using the UK Biobank Resource under Application Number 648, 56607, and 43017. We used the ALICE High Performance Computing Facility at the University of Leicester. The study was supported by Wellcome Trust Award WT225221/Z/22/Z. The study was partially supported by the NIHR Leicester Biomedical Research Centre and an NIHR Senior Investigator Award to M.D.T, and by an Asthma + Lung UK Career Development Award (AUK-CDA-2019-414); views expressed are those of the author(s) and not necessarily those of the NHS, the NIHR or the Department of Health. The funders had no role in the design of the study.

