## Supplementary Figures for "The role of the *ADRB2* Thr164Ile variant in lung function determination, plasma proteome variability and other phenotypes in UK Biobank"

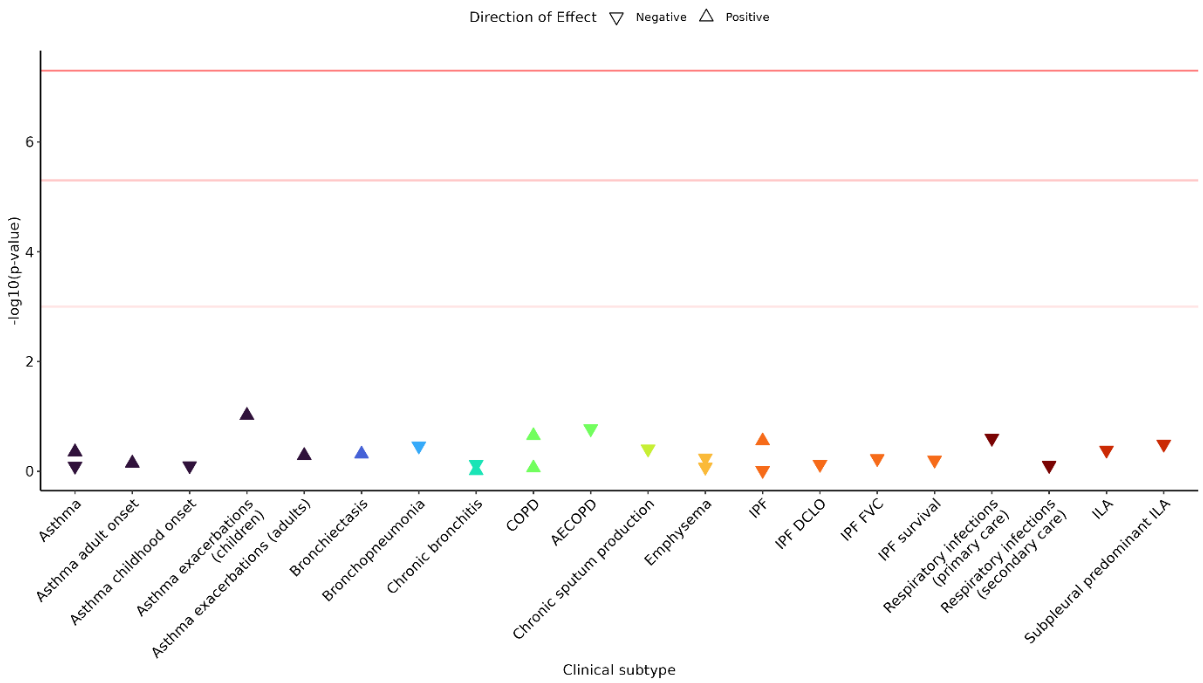


Supplementary Figure 1. Association of Thr164Ile variant (rs1800888) with respiratory diseases based on publicly available and in-house genome-wide association studies. Top to bottom, the three red lines represent the p value thresholds $5{\times}10^{-8}$, $5{\times}10^{-6}$ and $1{\times}10^{-3}$.

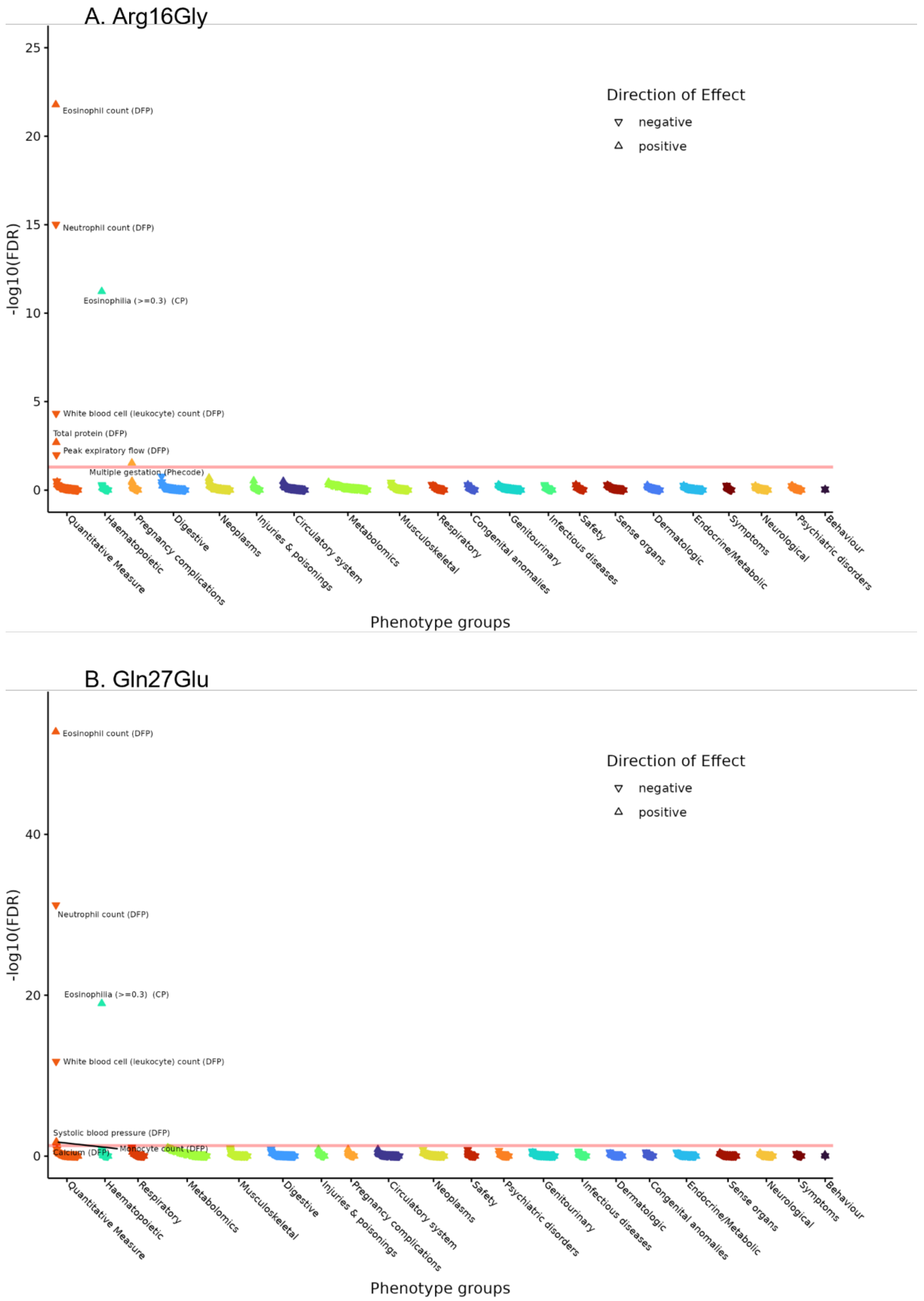


Supplementary Figure 2. Phenome-wide association of UK Biobank traits with Arg16Gly (A) and Gln27Glu (B) polymorphisms. Directions of effect are aligned to the minor A allele of Arg16Gly and the major C allele of Gln27Glu.

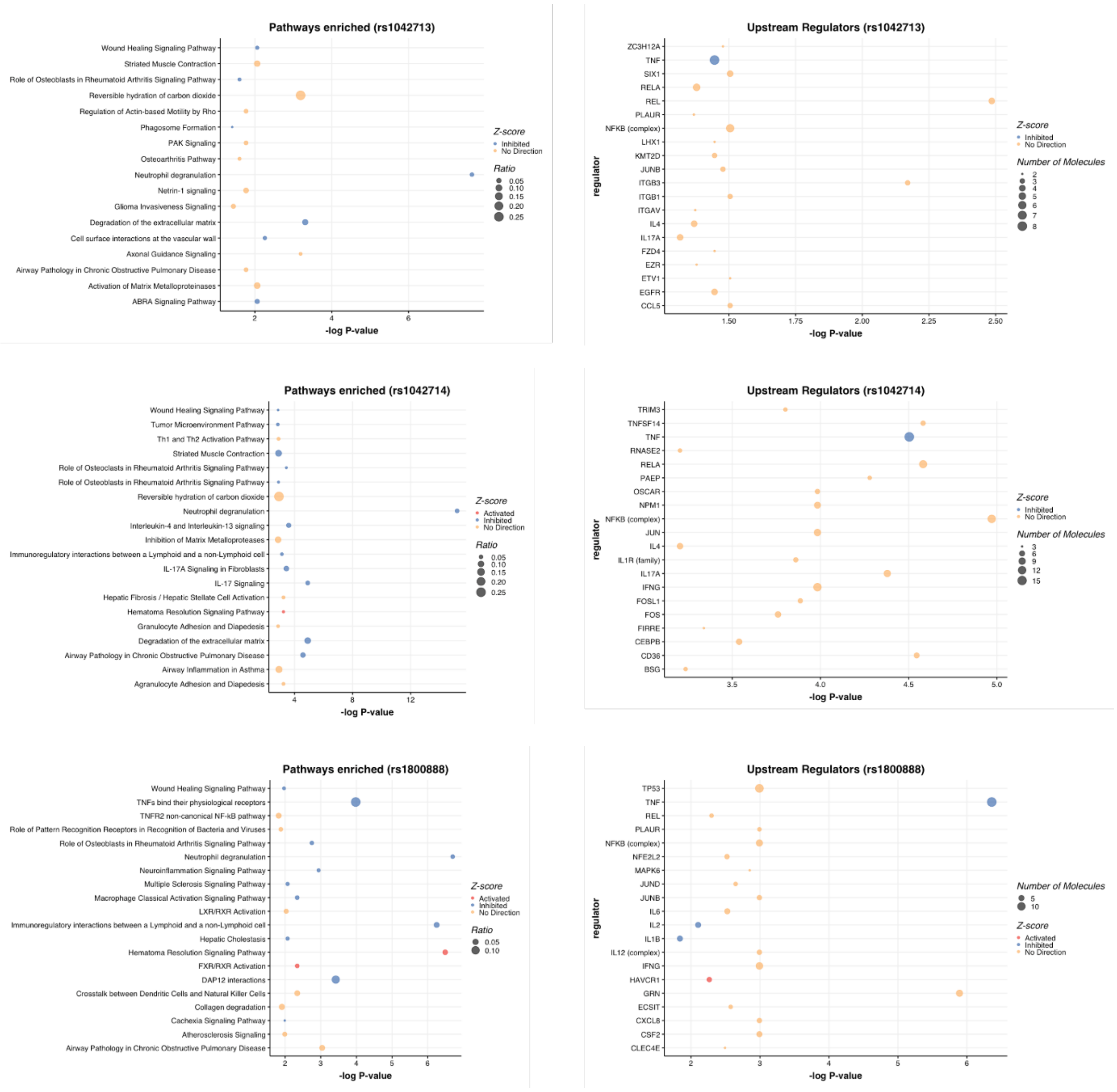


Supplementary Figure 3. Bubble plots of enriched pathways and upstream regulators amongst proteins associated with Arg16Gly (rs1042713), Gln27Glu (rs1042714) and Thr164Ile (rs1800888), generated using Ingenuity Pathway Analysis.

A

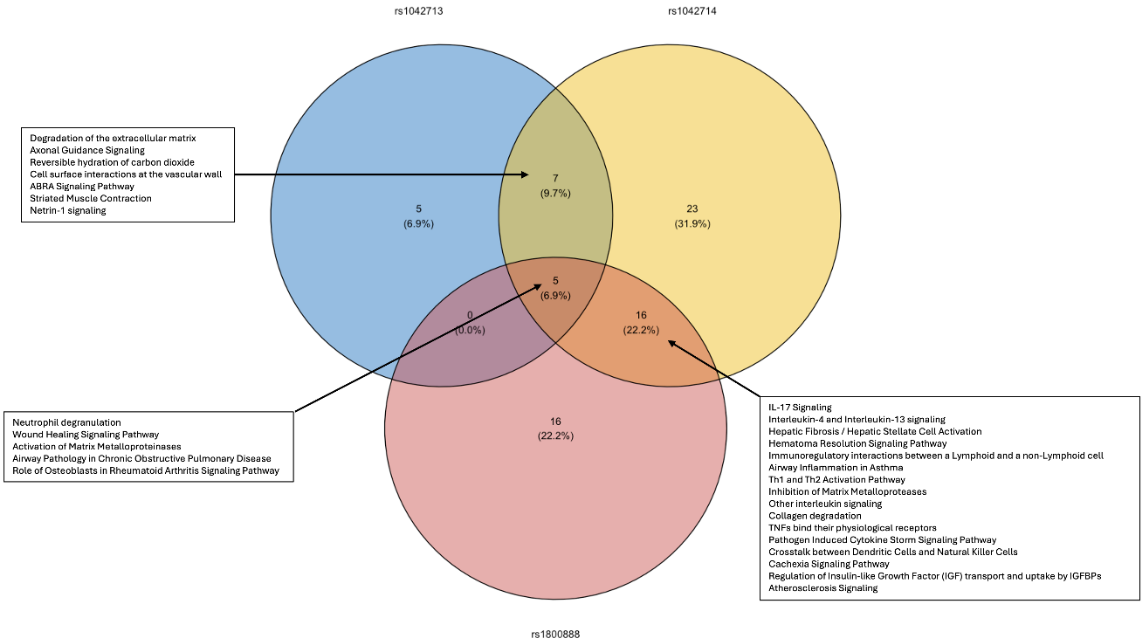


B

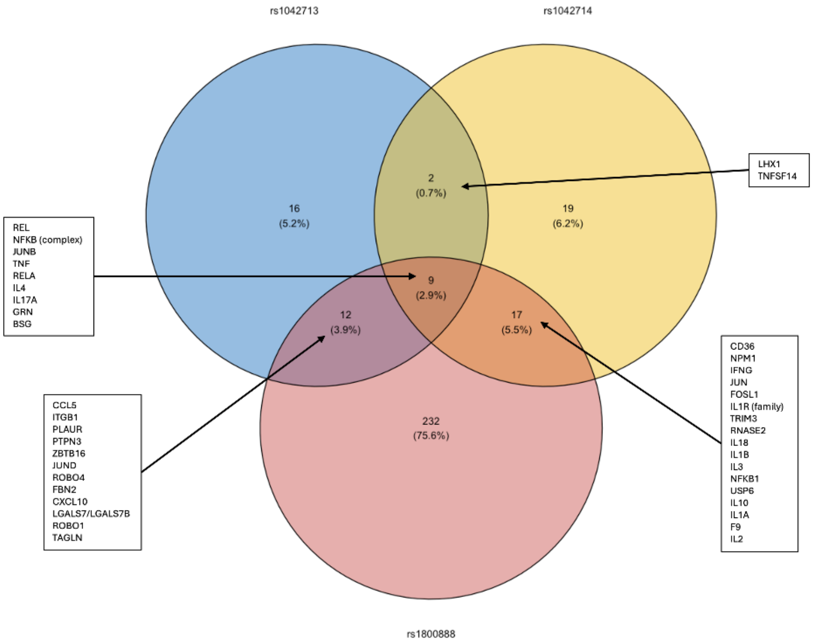


Supplementary Figure 4. Overlap of enriched canonical pathways (A) and upstream regulators (B) amongst proteins associated with Arg16Gly (rs1042713), Gln27Glu (rs1042714) and Thr164Ile (rs1800888).
